# Gastrointestinal carriage of *Klebsiella pneumoniae* in a general adult population in Norway: a cross-sectional study of risk factors and bacterial genomic diversity

**DOI:** 10.1101/2021.02.06.21251253

**Authors:** Niclas Raffelsberger, Marit Andrea Klokkhammer Hetland, Kristian Svendsen, Lars Småbrekke, Iren H. Löhr, Lotte Leonore Eivindsdatter Andreassen, Sylvain Brisse, Kathryn E. Holt, Arnfinn Sundsfjord, Ørjan Samuelsen, Kirsten Gravningen

## Abstract

**Background:** *Klebsiella pneumoniae* is a leading public health threat due to its increasing prevalence of antibiotic resistance. Gastrointestinal carriage of *K. pneumoniae* is a risk factor for subsequent infections in hospitalised patients. We determined risk factors for gastrointestinal carriage and the genomic population structure of *K. pneumoniae* colonising humans in a representative sample of a general population.

**Methods:** 2,975 individuals (54% women) ≥40y participating in the population-based Tromsø Study 7 (2015-2016) were included. Faecal samples were screened for *K. pneumoniae* which were characterised using whole-genome sequencing. Risk factors for carriage were analysed using data from the Norwegian Prescription Database and questionnaires, using multivariable logistic regression.

**Findings:** Prevalence of *K. pneumoniae* gastrointestinal carriage was 16·3% (95% CI 15·0-17·7%) with no gender difference. Risk factors associated with carriage included age ≥60y, travel to Greece or Asia past 12 months (adjusted odds ratio 1·49, 95% CI 1·11-2·00), Crohn’s disease/ulcerative colitis (2·26, 1·20-4·27), use of protein pump inhibitors (1·62, 1·18-2·22) and non-steroidal anti-inflammatory drugs past six months (1·38, 1·04-1·84), and antibiotic use last month (1·73, 1·05-2·86). Prevalence was higher among those having used combinations of drug classes and decreased over time with respect to preceding antibiotic use. The *K. pneumoniae* population was diverse with 300 sequence types among 484 isolates distributed across four phylogroups. Among the isolates, 5·2% and 11·6% harboured acquired resistance and virulence factors, respectively.

**Interpretation:** Identification of risk factors for gastrointestinal carriage in a representative sample of a general population allows for identification of individuals that may have a higher risk of extraintestinal infection during hospitalisation. The diverse population structure of *K. pneumoniae* carriage isolates reflects the ecologically adaptive capacity of the bacterium, and the low antibacterial consumption probably contributes to the low prevalence of resistance in clinical isolates in Norway.

## Introduction

*Klebsiella pneumoniae* (Kp) is a key pathogen associated with nosocomial infections frequently accompanied by antibiotic resistance.^1^ As an opportunistic pathogen, Kp is particularly problematic among neonates, elderly, immunocompromised, and patients with underlying chronic diseases, and commonly causes pneumonia, urinary tract infections, and bacteraemia.^1^ Additionally, the problem is exacerbated by the emergence and spread of community-acquired hypervirulent Kp causing infections in healthy individuals, usually presenting as pyogenic liver abscess occasionally accompanied with metastatic spread, but also as meningitis or endophthalmitis.^2^

Kp is known for its high prevalence and diversity of antibiotic resistance genes that challenge treatment options due to infections with multi-drug resistant variants. In Europe, antibiotic resistant Kp was responsible for more than 89,000 infections and 5,600 attributable deaths in 2015.^3^ Several new antibiotic resistance genes were first discovered within Kp before spreading to other pathogens.^4^ Consequently, The World Health Organisation considers antibiotic resistant Kp as a critical-priority bacterium in antibiotic research and development.^5^

Recent taxonomic updates show that Kp subdivides into five different species comprising seven phylogroups (Kp1-Kp7) referred to as the *K. pneumoniae* species complex (KpSC).^6^ The phylogroups include *K. pneumoniae sensu stricto* (Kp1), *K. quasipneumoniae* subsp. *quasipneumoniae* (Kp2), *K. variicola* subsp. *variicola* (Kp3), *K. quasipneumoniae* subsp. *similipneumoniae* (Kp4), *K. variicola* subsp. *tropica* (Kp5), *‘K. quasivariicola’* (Kp6) and *K. africana* (Kp7).^6^ Herein we refer to “Kp” for all members of the KpSC unless otherwise specified. Kp has a broad environmental distribution and transmission routes to humans are not defined.^6^

Gastrointestinal carriage of Kp as a reservoir for healthcare-associated Kp infections was established in the early 1970’s.^7^ Recent genomic studies show that gastrointestinal carriage is a risk factor for subsequent extraintestinal infections, and ∼50% of Kp bloodstream infections are caused by the patient’s own gut isolates.^8,9^ Moreover, the relative abundance of Kp in the gastrointestinal tract is associated with an increased risk of Kp bacteraemia.^10^

Cross-sectional studies have shown that Kp gastrointestinal carriage prevalence varies from 6% to 88% depending on geographical locations, detection methods and the populations investigated.^8,9,11,12^ However, we have a sparse understanding of risk factors for Kp gastrointestinal carriage and the population structure of Kp in the general human population. In a recent cross-sectional study of 911 pregnant women in low-income countries, Huynh et al. identified various country-specific environmental exposure factors linked to Kp gut carriage and a diverse Kp population structure.^12^

Here, we investigated the prevalence of Kp carriage and associated risk factors among 2,975 participants in a cross-sectional study of a representative sample of a general adult population in Norway, a country with a low prevalence of antibiotic resistance and restricted antibiotic use. Additionally, we elucidated the Kp genomic population structure of carriage isolates.

## SUBJECTS and METHODS

### Study population and design

The Tromsø study is a population-based study with repeated cross-sectional health surveys in the municipality of Tromsø, Norway. Tromsø is considered as representative of a Northern European, urban population.^13^ The seventh survey (Tromsø 7, https://uit.no/research/tromsostudy) was conducted March 2015-October 2016 and included two clinical visits. Unique national identity numbers from the official population-registry were used to invite all citizens ≥40 years (n=33,423) (Figure 1). Sixty-five percent (n=21,083, 11,074 women) attended the first clinical visit in the study. 9,324 attending the first visit, were invited for a second visit. These represent a random selection of 20% in age-group 40-59 years and 50% in age-group 60-84 years of the initially invited participants (n=33,423). To enhance the proportion of participants in earlier Tromsø Studies, 3,154 participants aged 40-84 years who had attended clinical examinations in Tromsø 6 were also invited. From March 2015 to March 2016, 5,800 of the 9,324 participants invited for the second visit were consecutively offered a faecal self-sampling kit. In total, 87% (n=5,042) returned a faecal sample either at the second visit, or by mail to the laboratory. Participants collected faecal material using nylon-flocked ESwab 490CE.A (Copan, Brescia, Italy). The first 3,009 of the 5,042 collected faecal samples were consecutively screened for the presence of Kp via selective culture. Due to resource limitations, the remaining 2,033 samples were not screened. All participants completed two self-administered structured questionnaires on socio-demographics, smoking, alcohol use, hospitalisation, chronic diseases, and travel abroad. After excluding 12 participants with wrong or missing sample identification number and 22 with missing questionnaires, our final study population consists of 2,975 participants.

**Figure 1:**
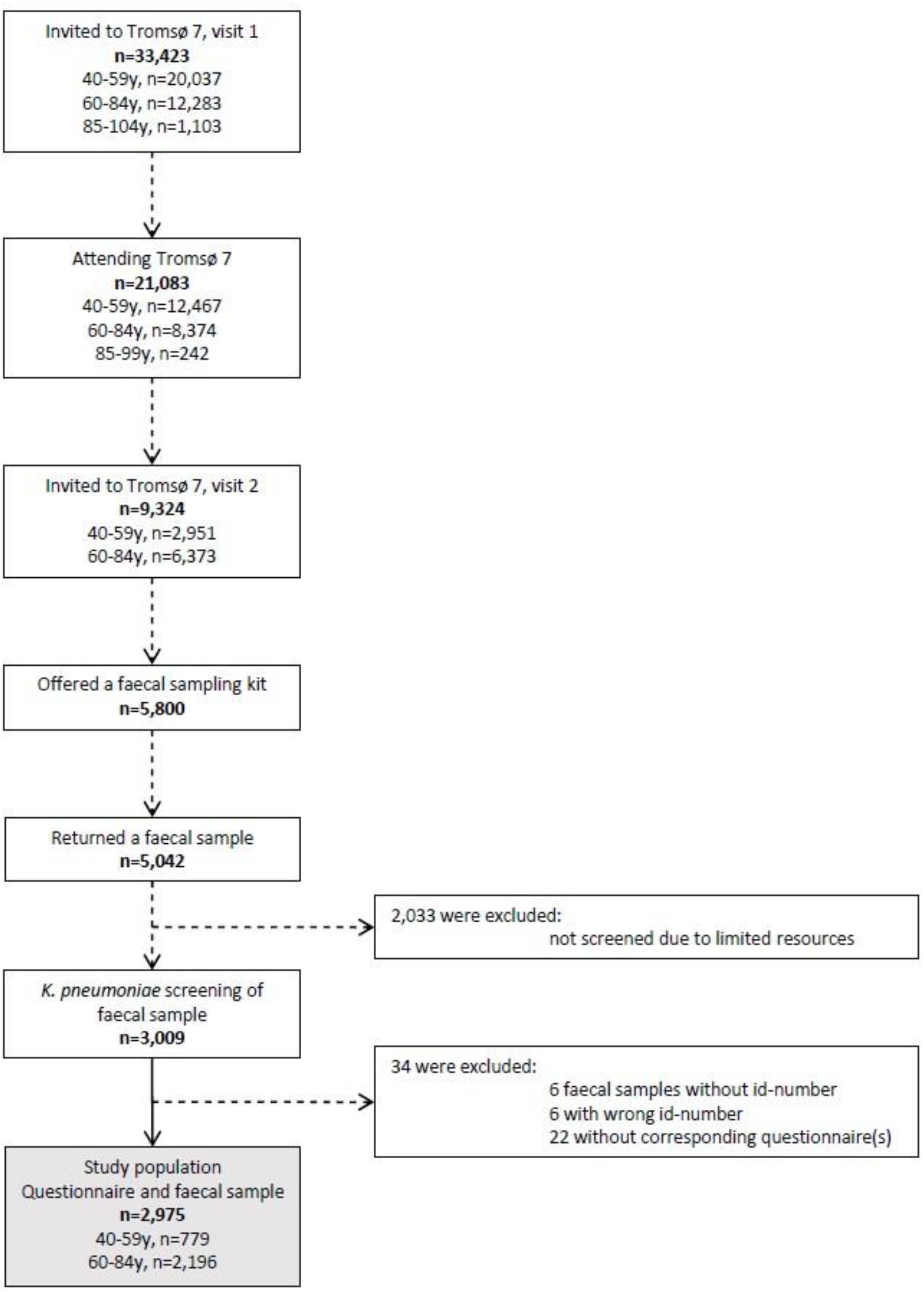
Flow diagram of study population

To assess the participants’ drug use during the preceding 12 months, data from Tromsø 7 were linked to the Norwegian Prescription Database (NorPD, http://www.norpd.no/). NorPD contains detailed information at the individual level on all dispensed prescription-drugs at all pharmacies in Norway. We defined drugs dispensed as drugs used and included the following groups in the Anatomical Therapeutic Chemical (ATC) classification system: A02 (acid related disorders), A10 (diabetes), H03A (thyroid hormones), J01, A07AA09, P01AB01 (antibacterials for systemic use), and M01 (anti-inflammatory and anti-rheumatic drugs).

### *Klebsiella pneumoniae* isolation

Upon arrival in the laboratory, 200 µl 85% glycerol was added to each ESwab tube and the samples were stored at −80°C. From the thawed media, 100 µl were plated onto Simmons citrate agar with inositol (SCAI) (both Sigma-Aldrich, Darmstadt, Germany) and incubated for 48 hours at 37°C.^14^ Large, yellow, glossy colonies suspected of being *Klebsiella* spp. were identified using mass spectrometry (MALDI-TOF, Bruker Daltonics, Bremen, Germany). The first colony identified as either *K. pneumoniae* or *K. variicola* from each sample was kept and further analysed. All samples were plated on cysteine lactose electrolyte deficient agar (MAST Group, Bootle, UK) to assess growth of faecal flora and validity of the samples.

### Antimicrobial susceptibility testing

Susceptibility testing was performed according to the EUCAST disk diffusion method and interpreted using the EUCAST 2021 breakpoint table (https://eucast.org/).

### Genomic sequencing and bioinformatic analyses

DNA was extracted with the MagNA Pure 96 system (Roche Applied Science, Manheim, Germany) and sequencing libraries were prepared according to the Nextera Flex sample preparation protocol (Illumina, San Diego, CA, USA). Sequencing was performed on the Illumina MiSeq platform to generate 300 bp paired-end reads. All reads were assembled with Unicycler v0.4.8.^15^ Kleborate v2.0.0 was used to determine sequence type (ST), species identification, and acquired genes encoding virulence or antibiotic resistance.^16^ Kaptive was used to identify capsule biosynthesis loci (KL), and LPS (O) antigen loci. Plasmid replicons were identified with PlasmidFinder v2020-07-13 using Abricate v0.9.9 (https://github.com/tseemann/abricate). Novel STs, *bla*_SHV_, *bla*_LEN_, *bla*_OKP_, and virulence alleles were assigned by the curators of the Institut Pasteur multilocus sequence type (MLST) and core genome MLST databases (https://bigsdb.pasteur.fr/klebsiella). To verify the absence of *bla*_SHV_/*bla*_LEN_, observed in two genomes, the raw reads were inspected with SRST2 v0.2.0.^17^

### Phylogenetic analyses

To assess the phylogenetic relatedness, a core genome alignment of the 484 genomes against the *K. pneumoniae* ST23 NTUH-K2044 reference chromosome (GenBank accession: NC_012731.1) was generated using the RedDog pipeline v1beta.11 (https://github.com/katholt/RedDog), and inferred as described previously.^18^ To identify the number of single-nucleotide polymorphisms (SNPs) between any two genomes from the resulting alignment, snp-dists v0.7.0 (https://github.com/tseemann/snp-dists/) was used.

### Definition of hypervirulent Kp

Hypervirulent Kp were defined here according to Huynh *et al*., as isolates harbouring at least one of the genes *rmpA* and *rmpA2*, and/or at least one complete gene cluster among *iucABCD*-*iutA* (aerobactin) and *iroBCDN* (salmochelin).^12^

### Statistical analyses

The primary analysis was a multivariable logistic regression model with outcome variable being culture-confirmed Kp gastrointestinal carriage using SPSS v26.0 (SPSS, Inc., Chicago, IL, USA). Explanatory variables were selected with the help of a directed acyclic graph constructed using DAGitty v3.0 (Suppl. Fig. 1).^19^ All explanatory variables were kept in the fully adjusted model. The strength of the associations was examined by calculating adjusted odds ratios (AORs) with 95% confidence intervals (CIs). Two-sided p-values <0.05 were considered statistically significant. We used R v4.0.0 (Foundation for Statistical Computing, Vienna, Austria) to create a proportional Venn diagram for statistically significant drug groups associated with Kp carriage. STATA v16.1 (StataCorp LLC, Texas, USA) was used to analyse the cumulative change in proportion of Kp carriers associated with antibiotic use in the past 1-12 months.

### Ethics

The study, including the linking of data between Tromsø 7 and NorPD, was approved by the Regional Committee for Medical and Health Research Ethics, North Norway (REC North reference: 2016/1788 and 2014/940) and the Data Protection Officer at University Hospital of North Norway (reference: 2019/4264). The study complied with the Declaration of Helsinki. All participants in Tromsø 7 signed an informed consent form prior to participation.

## RESULTS

We analysed data from 2,975 participants (1,615 women, 54·3%, Suppl. Table 1). Median age of participants was 65·0 years (Interquartile Range 59-71 years, no gender difference). Altogether, we identified 484 Kp carriers corresponding to a prevalence of 16·2% (95% CI 14·5-18·1) among women and 16·3% (14·4-18·4) among men.

**Table 1.** *K. pneumoniae* (Kp) faecal carriage and associated factors among 2,975 participants in Tromsø 7.

|  | % (Kp) | n (Kp) | Denominator | AOR‡ | 95% CI | p-value |
| --- | --- | --- | --- | --- | --- | --- |
| **Age (years)** |  |  |  |  |  | 0.128 |
| 40-49 | 10.8 | 37 | 344 | 1.00 |  |  |
| 50-59 | 15.4 | 67 | 435 | 1.34 | 0.85-2.11 |  |
| 60-69 | 17.3 | 222 | 1,286 | 1.56 | 1.06-2.30 |  |
| 70-84 | 17.4 | 158 | 910 | 1.56 | 1.03-2.36 |  |
| **Current daily smoking** |  |  |  |  |  | 0.717 |
| No | 16.2 | 421 | 2,598 | 1.00 |  |  |
| Yes | 15.7 | 55 | 351 | 0.94 | 0.66-1.34 |  |
| **Alcohol consumption frequency** |  |  |  |  |  | 0.050 |
| Never to ≤ monthly | 16.4 | 164 | 998 | 1.00 |  |  |
| 2-4/month to 2-3/week | 16.9 | 298 | 1,765 | 1.13 | 0.88-1.46 |  |
| ≥4/week | 9.1 | 18 | 198 | 0.61 | 0.35-1.05 |  |
| **Alcohol units/occasion** |  |  |  |  |  | 0.365 |
| 1-4 | 16.2 | 412 | 2,540 | 1.00 |  |  |
| ≥5 | 14.1 | 20 | 142 | 0.79 | 0.47-1.32 |  |
| **Travel abroad past 12 months<sup>a</sup>** |  |  |  |  |  | 0.009 |
| No | 15.5 | 196 | 1,276 | 1.00 |  |  |
| Greece or Asia | 20.3 | 102 | 502 | 1.49 | 1.11-2.00 |  |
| All other countries | 15.3 | 179 | 1,171 | 0.97 | 0.76-1.26 |  |
| **Hospitalisation last 12 months** |  |  |  |  |  | 0.844 |
| No | 15.9 | 412 | 2,588 | 1.00 |  |  |
| Yes | 19.0 | 67 | 353 | 1.03 | 0.74-1.44 |  |
| **Diabetes mellitus<sup>b</sup>** |  |  |  |  |  | 0.056 |
| No | 16.0 | 427 | 2,674 | 1.00 |  |  |
| Yes | 23.5 | 40 | 170 | 1.82 | 0.99-3.36 |  |
| **Crohn's disease/ulcerative colitis** |  |  |  |  |  | 0.012 |
| No | 16.0 | 452 | 2,831 | 1.00 |  |  |
| Yes | 28.3 | 17 | 60 | 2.26 | 1.20-4.27 |  |
| **Proton pump inhibitors last 6 m<sup>c</sup>** |  |  |  |  |  | 0.003 |
| No | 15.3 | 404 | 2,632 | 1.00 |  |  |
| Yes | 23.3 | 80 | 343 | 1.62 | 1.18-2.22 |  |
| **NSAIDs last 6 months<sup>d</sup>** |  |  |  |  |  | 0.028 |
| No | 15.6 | 397 | 2,545 | 1.00 |  |  |
| Yes | 20.2 | 87 | 430 | 1.38 | 1.04-1.84 |  |
| **Antibiotic systemic use last 1 m<sup>e</sup>** |  |  |  |  |  | 0.032 |
| No | 15.8 | 453 | 2,866 | 1.00 |  |  |
| Yes | 28.4 | 31 | 109 | 1.73 | 1.05-2.86 |  |
| **Metformin last 6 months<sup>f</sup>** |  |  |  |  |  | 0.764 |
| No | 16.0 | 460 | 2,867 | 1.00 |  |  |
| Yes | 22.2 | 24 | 108 | 0.89 | 0.40-1.95 |  |
| **Thyroid hormones last 6 months<sup>g</sup>** |  |  |  |  |  | 0.333 |
| No | 15.9 | 431 | 2,714 | 1.00 |  |  |
| Yes | 20.3 | 53 | 261 | 1.20 | 0.83-1.75 |  |

‡AOR, adjusted odds ratio; CI, confidence interval; m, months; NSAIDs, non-steroidal anti-inflammatory drugs; NorPD, Norwegian Prescription Database. AOR adjusted for age, current daily smoking, alcohol consumption frequency, alcohol units/occasion, travel abroad past 12 months, hospitalisation last 12 months, diabetes mellitus, Crohn's disease/ulcerative colitis and drug use according to NorPD (A02BC, M01, J01, A07AA09, P01AB01, A10BA02, H03AA). The multivariable model contains 2,446 participants with complete information on all variables.
<sup>a</sup> Travelled outside the Nordic countries >1 week duration in the past 12 months.
<sup>b</sup> 20 participants who answered "Yes, previously" were excluded.
<sup>c</sup> A02BC, drugs used for peptic ulcer and gastro-oesophageal reflux disease.
<sup>d</sup> M01, anti-inflammatory and anti-rheumatic products (non-steroids), anti-inflammatory/anti-rheumatic agents in combination and specific anti-rheumatic agents.
<sup>e</sup> J01, A07AA09, P01AB01, antibacterials for systemic use, intestinal anti-infectives and nitroimidazole derivatives used as antiprotozoals (metronidazole).
<sup>f</sup> A10BA02, blood glucose lowering drug used in diabetes.
<sup>g</sup> H03AA, natural and synthetic thyroid hormones.

### Kp faecal carriage and associated factors

In analyses adjusting for all of the explanatory variables, Kp faecal carriage was associated with age 60 years and older, self-reported travel to Greece or Asia during the preceding 12 months (AOR 1·49, 1·11-2·00) and Crohn’s disease/ulcerative colitis (2·26, 1·20-4·27) (Table 1). Furthermore, carriage was associated with the use of proton pump inhibitors (PPIs) (1·62, 1·18-2·22) and non-steroidal anti-inflammatory drugs (NSAIDs) (1·38, 1·04-1·84) in the last six months and antibiotic use in the last month (1·73, 1·05-2·86). Analysis of the significant variables in the multivariable model for the three most frequent species is presented in Suppl. Table 2.

The Kp prevalence of the three statistically significantly associated drug classes (usage in past six months) in relation to Kp carriage is shown in Figure 2. Kp prevalence was 16·1% among 286 antibiotic-only users, 17·4% among 305 NSAID-only users and 21·5% among 209 PPI-only users. Kp prevalence increased in each overlapping area for two drug classes, and further increased to 30·8% in the overlapping area for all three.

**Figure 2.**
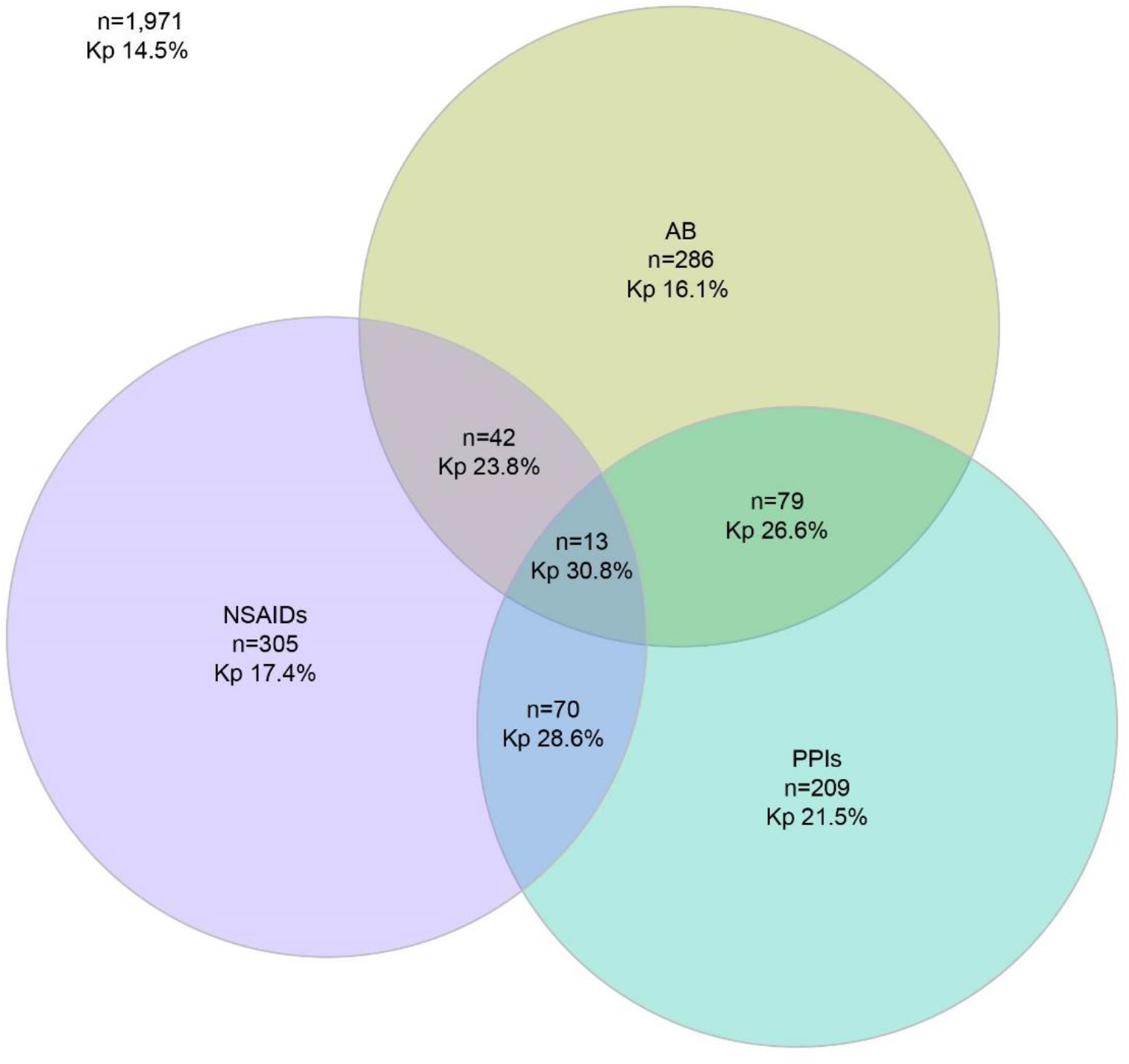
Proportional Venn diagram of Kp carriage prevalence related to statistically significantly associated drug classes (antibiotics (AB), nonsteroidal anti-inflammatory drugs (NSAIDs) and proton pump inhibitors (PPIs)) the past 6 months among 2,975 study participants.

Looking at the cumulative change in proportion of Kp carriers associated with antibiotic use during 1-12 months before faecal sampling, we found that Kp carriage prevalence was highest amongst those with antibiotic use in the past month (28·4%), or past two months (25·0%), decreasing to around 20·0% in the past 6-12 months (Figure 3). In contrast, in the non-antibiotic using population, prevalence of Kp carriage was significantly lower (15·2%).

**Figure 3:**
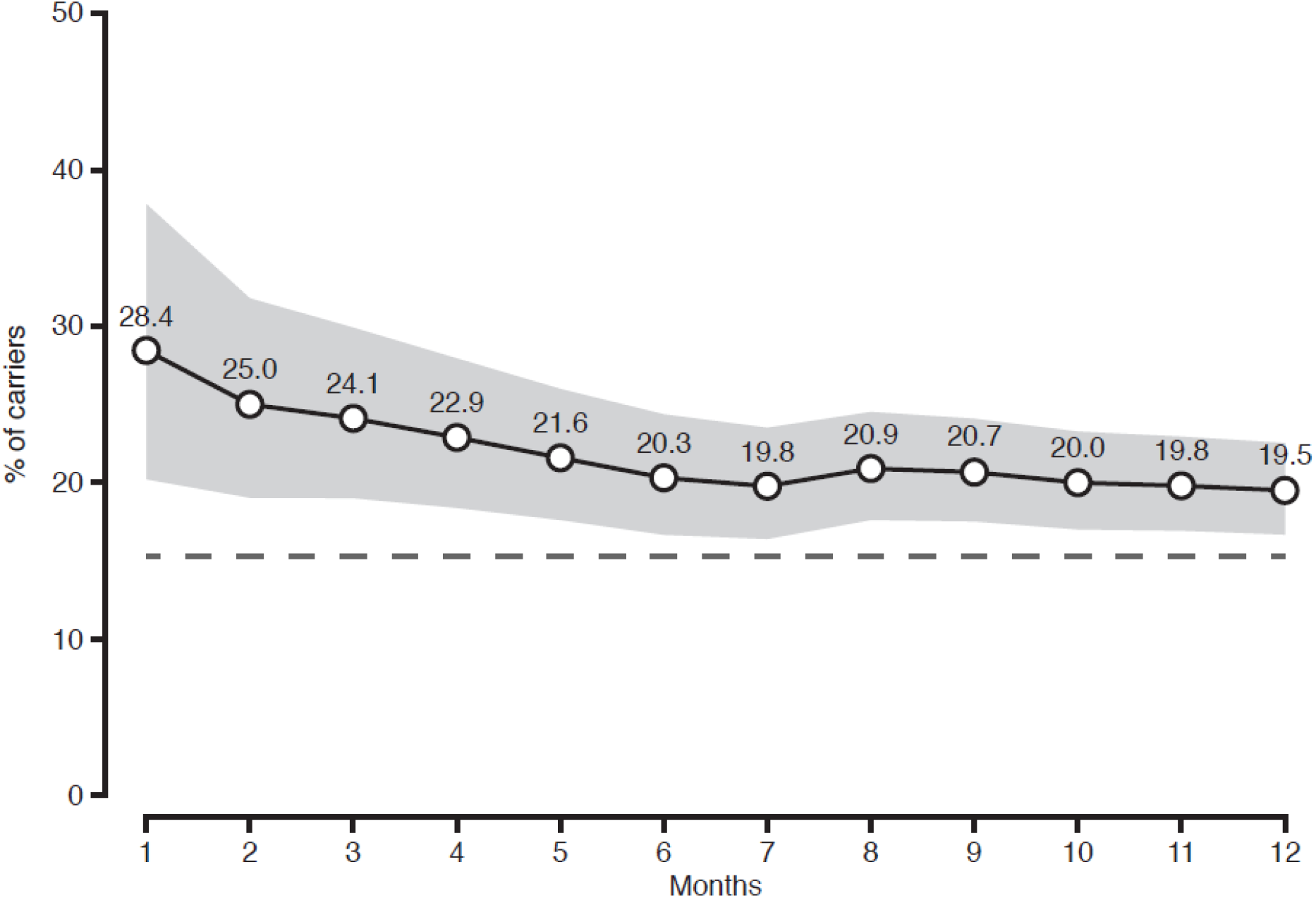
Cumulative change in the proportion of Kp carriers among those who had used antibiotics 1-12 months before the faecal sampling. Shaded grey area represents the 95% CI. The time period at each specified month includes data for the preceding months. The dashed black line indicates the prevalence of carriage in the non-antibiotic using population (15·2%).

### Kp phylogeny and diversity

Phylogenetic analysis based on whole-genome sequencing of 484 *K. pneumoniae* isolates identified three species distributed in four phylogroups with a predominance of *K. pneumoniae sensu stricto* (Kp1, 62·6%), followed by *K. variicola* subsp. *variicola* (Kp3, 27·7%), *K. quasipneumoniae* subsp. *quasipneumoniae* (Kp2, 6·4%), and *K. quasipneumoniae* subsp. *similipneumoniae* (Kp4, 3·3%) (Figure 4, Suppl. Table 3). Phylogroups Kp5-Kp7 were not identified. Using MLST, we found a high degree of genotypic diversity (Simpson diversity index 99·5%) with a total of 300 different STs, including 96 (32%) novel STs (Suppl. Fig. 2). The majority (79%) of the STs were represented by a single isolate. Only 17 STs (5·7%) were represented by more than five isolates. The most frequent were ST20 (n=15, 3·1%), ST26 (n=13, 2·7%), ST35 (n=9, 1·9%), ST37 (n=9, 1·9%), and ST2386 (n=9, 1·9%). With regard to clonal relatedness we found seven ST35 and three ST25 isolates with a range of zero to five SNPs (Suppl. Table 4), and seven to eight SNPs (Suppl. Table 5), respectively, indicating a recent clonal spread. No close clonal relatedness was identified among the remaining frequent STs (Suppl. Table 6).

**Figure 4:**
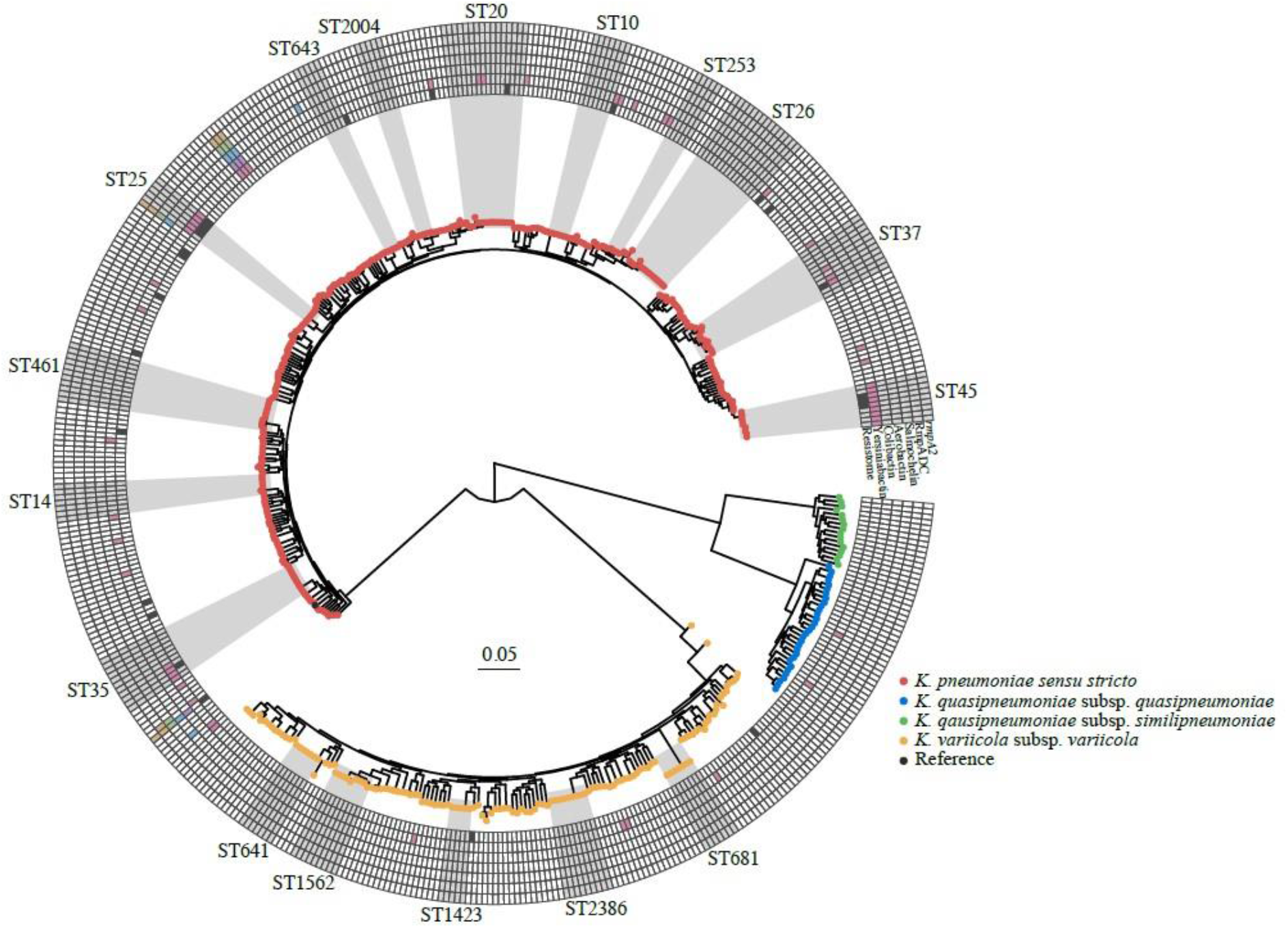
Core chromosomal maximum likelihood phylogeny of the 484 Kp genomes. The tips are coloured by species. The heatmap shows presence (colour) or absence (white) of acquired resistance genes (innermost ring) or virulence factors (remaining six rings). Clades corresponding to STs with five or more genomes are highlighted and labelled.

### Antimicrobial resistance and plasmid content

The prevalence of resistance was low and none of the isolates were resistant to cefotaxime, meropenem, aztreonam, or ciprofloxacin (Suppl. Fig. 3). Resistance was observed against amoxicillin-clavulanic acid (0·6%), ceftazidime (0·2%), gentamicin (0·4%), cefuroxime (0·6%), trimethoprim-sulfamethoxazole (1·7%), piperacillin-tazobactam (4·8%), and mecillinam (5·0%). This was in concordance with the low number of intrinsic and acquired resistance genes (Figure 4, Suppl. Table 3). A high sequence diversity of species-specific intrinsic narrow-spectrum chromosomal β-lactamase gene alleles were identified (Suppl. Fig. 4, 5, 6, and 7, Suppl. Table 3). Three ampicillin susceptible isolates harboured either deleted *bla*_SHV_/*bla*_LEN_ genes or a premature stop codon in *bla*_SHV_. Only 5·2% of the isolates harboured an acquired resistance gene (median number: 2, Suppl. Table 3). *Bla*_SHV-51_ and *bla*_SHV-52_ were assigned by Kleborate as extended-spectrum β-lactamase and inhibitor resistant β-lactamase, respectively. However, both isolates were phenotypically susceptible to all tested β-lactams and β-lactam-inhibitor combinations. Thirty-two plasmid replicon types were identified among 393/484 (81·2%) isolates with IncFIB(K) (25·2%), Col(pHAD28) (15·0%), IncFIB(K) (pCAV1099-114) (11·4%), IncFIA(HI1) (10·5%), and IncFII(pKP91) (9·8%) the most frequent (Suppl. Table 3). The median number of replicon types per isolate was two.

### Virulence factors and serotype prediction

The majority of isolates (n=428, 88·4%) did not contain known acquired virulence determinants (Suppl. Table 3). Five *K. pneumoniae sensu stricto* isolates (1·0%) of different STs were defined as hypervirulent, including one isolate of the known hypervirulent ST23 clone.^2^ These five isolates harboured aerobactin (*iuc1* or *iuc2*), salmochelin (*iro1* or *iro2*) and *rmpA* (Suppl. Table 3); three additionally carried the *ybt* locus. Three of the hypervirulent isolates harboured the capsular synthesis locus (KL) 1 or KL2, previously described as hypervirulence associated^2^; the other two harboured KL23. Overall, the siderophore loci *ybt* (yersiniabactin), *iuc* (aerobactin), and *iro* (salmochelin) were present in 10·7% (n=52), 1·4% (n=7), and 1·0% (n=5) of the isolates, respectively (Suppl. Table 3). A total of 96 defined KL types were found (Suppl. Fig. 8), with KL10 being the most frequent (n=21; 4·7%) followed by KL28 (n=18; 4·0%) and KL22 (n=17; 3·8%). KL2 accounted for 2·9% (n=13) and KL1 for only 0·9% (n=4). We observed 11 different defined O-antigen types in 477 isolates (Suppl. Fig. 9) with O1 (n=133; 27·9%) being the most frequent followed by O3/3a (n=113; 23·7%), O2 (n=76; 15·9%), O5 (n=58; 12·2%), and O3b (n=47; 9·9%), accounting for 90% of the isolates. Five isolates harboured an O-locus with an undefined O-type.

## Discussion

In this large study of a representative sample of a general population in Norway, we detected an overall Kp faecal carriage prevalence of 16.3% with no gender difference. We found that Kp carriage was associated with age 60 years and older, reported Crohn’s disease/ulcerative colitis, travel to Asia and Greece in the preceding 12 months, and recent use of PPIs, NSAIDs, and antibiotics. We showed that the Kp population among adults in the community setting in a high-income country with low antibiotic use was highly diverse and characterised by a low prevalence of acquired antibiotic resistance and virulence determinants.

The identified Kp carriage prevalence of 16·3% is lower than that observed among pregnant women in low-income countries (40-66%)^12^ and among healthy adults in Asian countries (19-88%)^11^, but in range with a hospital admission study in Australia (6-19%)^8^ and hospitalised patients in the USA (23%).^9^ This could be explained by the different populations investigated (e.g. healthy individuals, hospitalised patient groups, geographic setting, and/or ethnicity), but also by the sampling and detection strategies. Consistent with the culture-based Australian study of hospitalised patients on admission, we found that Kp carrier prevalence increased with age.^8^ The association of Kp gastrointestinal carriage with travel abroad, irrespective of resistance phenotype, has to our knowledge not been demonstrated before. The higher Kp prevalence associated with travel to Asia could be explained by the observed high carriage among people in this region ^11,12^.

The increased prevalence of Kp carriage associated with Crohn’s disease/ulcerative colitis could be due to disease specific gut microbiota alterations. The gut microbiota of patients with Crohn’s disease departs from the normal state as microbial diversity is significantly diminished including an increased abundance of *Enterobacterales*.^20^ A review by Kaur et al. 2018 emphasises the possible role of Kp in the pathogenesis of lower intestinal tract diseases.^21^ This corresponds to our findings of a higher Kp carrier prevalence among participants with self-reported Crohn’s disease/ulcerative colitis. However, the pathogenesis of Crohn’s disease/ulcerative colitis is complex and involves an interplay between different factors which may include the microbiota composition.^22^

A large number of non-antibiotic drugs have been shown to inhibit growth of one or more representative bacterial species in the human gastrointestinal tract.^23^ The positive association of PPI and/or NSAID use with Kp carriage further underpins the influence of non-antibiotic drugs on faecal microbiota and the selection of specific bacterial species. PPI use is implicated in altered gut microbiota composition, bacterial colonisation patterns, including multi-drug resistant microorganisms, and increased susceptibility to enteric bacterial infections.^24,25^ Although no direct evidence exists, it may be possible that Kp as a major etiological agent of liver abscess^26^, particularly in Asian countries, could be linked to the observation that PPI therapy is associated with an increased risk of cryptogenic liver abscess.^27^ The role of NSAIDs as a risk factor is unclear, but NSAID use has been shown to influence the gastrointestinal microbiota towards a higher relative abundance of *Enterobacteriaceae*.^28^

Systemic antibiotics more obviously influence the microbiota composition. Kp is intrinsically resistant to amino-penicillins and carboxypenicillins, and thus has a selective advantage compared to other bacteria during treatment with penicillins, which constitutes approximately half of human antibiotic use in Norway.^29^ Antibiotics leave an imprint on the gastrointestinal bacterial community after treatment is removed, ranging from weeks to years in different studies.^30^ This is consistent with our finding of a significant increase in Kp prevalence related to antibiotic exposure even 12 months prior to faecal sampling. Notably, our data display a quantitative time-response relationship between antibiotic use and Kp carriage. The prevalence of Kp carriage decreased amongst individuals sampled from one to six months post-exposure and reached a plateau at 5% above that of the non-antibiotic using population sampled at 6-12 months post-exposure. The latter indicates a potentially long-lasting effect on faecal Kp occurrence after antibiotic exposure.

Comparable to previous reports on Kp carriage isolates in pregnant women in the community-setting in low-income countries^12^ and hospitalised patients in high-income countries^8,9^, we found a phylogenetically highly diverse Kp population with dominance of *K. pneumoniae sensu stricto*. This indicates individually adapted Kp populations with limited interconnection. The observed diversity in our study is also probably an underestimation due to the selection of only one colony for sequencing. Considering the evidence that a high proportion of Kp extraintestinal infections originate from the patients’ own carriage isolates^8,9^, we assume that the Kp carriage population structure among individuals in the community is partly mirrored in the clinical setting. This is in line with the finding that six out of the ten most frequent STs of the carriage isolates also are among the ten most frequently observed STs in an ongoing Kp bacteraemia study in Norway (unpublished data; Fostervold et al.). The low level of antibiotic resistance in our study is consistent with national data in the clinical setting and reflects the relatively low antibiotic consumption in Norway.^29^

This is the first study investigating the prevalence of Kp faecal carriage in a large representative sample of a general population in a community setting in a high-income country. Important methodological strengths include high study attendance rate. We used data from nearly 3,000 people aged ≥ 40 years, thereby avoiding the substantial selection bias related to convenience samples in healthcare settings. Major strengths also include the combined use of questionnaire data, drugs dispended at pharmacies from a national registry database, and both phenotypic and genomic laboratory results.

The age restriction is a limitation. It would have been interesting to analyse Kp prevalence among those below 40 years whom use less antibiotics and have fewer chronic diseases.^29^ As we found that Kp prevalence increased with age, extrapolation would suggest younger adults to have a lower prevalence. Due to lack of resources, we did not screen all available faecal samples for Kp carriage which would have further increased the precision of the estimates. We used selective SCAI medium for detection of Kp, as this strategy has been shown to have a high Kp recovery supporting the growth of all Kp phylogroups.^14^ However, we acknowledge that positive culturing could reflects those with a relative high abundance of Kp and that molecular based approaches, which are less dependent on abundance and phenotypic differentiation, may detect an even higher prevalence of Kp carriage. Home sampling, as conducted in our study, could be another biasing factor. However, we controlled for this by ensuring validity of the samples by assessing bacterial growth consistent with faecal flora on non-selective media and mean transport time from sampling to laboratory arrival was only 1·8 days.

In conclusion, our findings illustrate the association of non-antibiotic drugs and inflammatory bowel diseases with increased prevalence of Kp gastrointestinal carriage that warrants considerations with regard to risk stratification in the prevention of healthcare-associated infections. The highly diverse population structure of Kp colonising humans illustrates the capacity for adaptive diversification of this species complex. This complicates the identification of potential Kp cross-niche transmission, which will be important in detection of animal or environmental Kp reservoirs for clinically-relevant human Kp carriage and infection from a One Health perspective.

## Data Availability

Bacterial genome data (raw Illumina reads) are publicly available in NCBI under BioProject PRJEB42350. This study is based on data owned by a third party (The Tromsø Study, Department of Community Medicine, UiT The Arctic University of Norway). Confidentiality requirements according to Norwegian law prevents sharing of individual patient level data in public repositories. Application of legal basis and exemption from professional secrecy requirements for the use of personal health data in research may be sent to a regional committee for medical and health research ethics (https://helseforskning.etikkom.no). The authors gained access to the data through the Tromsø Study's application process. Guidelines on how to access the data are available at the website https://uit.no/research/tromsostudy. All enquiries about the Tromsø Study should be sent by e-mail to. All the questionnaire variables are published in the NESSTAR program system and results can be viewed online: https://uit.no/research/tromsostudy.

## Author contributions

ØS conceptualised and acquired funding for the study in collaboration with AS, KG, IHL, SB and KH. ØS was responsible for organising the collection of faecal samples. LLEA did the screening of faecal samples and phenotypic testing. NR, KS, LS and KG conceptualised and conducted the epidemiological analyses of risk factors. IHL organised whole-genome sequencing of isolates. MAKH, NR and ØS did the analysis of whole genome sequence data. SB and KH provided tools for genomic analysis and data curation. NR, MAKH, ØS, and KG prepared first manuscript draft. All authors contributed to review and editing of the manuscript and approved the final version.

## Acknowledgements

We are grateful for technical assistance from Bjørg Haldorsen, Bettina Aasnæs and Ellen Josefsen in organising the collection of faecal samples in the laboratory. Eva Bernhoff and Ragna-Johanne Bakksjø for performing whole-genome sequencing. Dorota Buczek for creating the Venn diagram. Rod Wolstenholme for figure editing. We thank the team of curators of the Institut Pasteur MLST and whole-genome MLST databases for curating the data and making them publicly available at https://bigsdb.pasteur.fr/.

## Data Availability

Bacterial genome data (raw Illumina reads) are publicly available in NCBI under BioProject PRJEB42350. This study is based on data owned by a third party (The Tromsø Study, Department of Community Medicine, UiT The Arctic University of Norway). Confidentiality requirements according to Norwegian law prevents sharing of individual patient level data in public repositories. Application of legal basis and exemption from professional secrecy requirements for the use of personal health data in research may be sent to a regional committee for medical and health research ethics (https://helseforskning.etikkom.no). The authors gained access to the data through the Tromsø Study’s application process. Guidelines on how to access the data are available at the website https://uit.no/research/tromsostudy. All enquiries about the Tromsø Study should be sent by e-mail to. All the questionnaire variables are published in the NESSTAR program system and results can be viewed online: https://uit.no/research/tromsostudy.

