## Supplementary Material for "Gastrointestinal carriage of *Klebsiella pneumoniae* in a general adult population in Norway: a cross-sectional study of risk factors and bacterial genomic diversity"

**Suppl. material.**

**Table of contents**

Supplementary Table 1. Characteristics for the study population, 2975 participants in Tromsø 7

Supplementary Table 2. Analysis of the significant variables in the multivariable model according to *Klebsiella* species

Supplementary Table 3. Genome characteristics of 484 *K. pneumoniae* species complex isolates (provided as Excel-file)

Supplementary Table 4. SNP matrix of ST35 isolates

Supplementary Table 5. SNP matrix of ST25 isolates

Supplementary Table 6. SNP distances within sequence types with more or equal than five isolates

Supplementary Figure 1. Directed acyclic graph (DAG)

Supplementary Figure 2. Multilocus sequence type (MLST) diversity

Supplementary Figure 3. Prevalence of phenotypic antimicrobial non-susceptibility

Supplementary Figure 4. SHV β-lactamase diversity among *K. pneumoniae sensu stricto* isolates

Supplementary Figure 5. LEN β-lactamase diversity among *K. variicola* subsp. *variicola* isolates

Supplementary Figure 6. OKP-A β-lactamase diversity among *K. quasipneumoniae* subsp. *quasipneumoniae* isolates

Supplementary Figure 7. OKP-B β-lactamase diversity among *K. quasipneumoniae* subsp. *similipneumoniae* isolates

Supplementary Figure 8: Capsule locus (KL) type diversity

Supplementary Figure 9. LPS (O) type diversity

**Supplementary Table 1.** Characteristics for the study population, 2,975 participants in Tromsø 7

| Characteristics | N | % |
|---|---|---|
| **Sex** | | |
| Men | 1,360 | 45·7 |
| Women | 1,615 | 54·3 |
| **Age** (years) | | |
| 40-49 | 344 | 11·6 |
| 50-59 | 435 | 14·6 |
| 60-69 | 1,286 | 43·2 |
| 70-84 | 910 | 30·6 |
| **Living with a spouse/partner** | | |
| No | 676 | 24·1 |
| Yes | 2,124 | 75·9 |
| **Education level** | | |
| Low | 1,745 | 60·0 |
| High[a] | 1,163 | 40·0 |
| **Household income** | | |
| Low | 1,250 | 44·6 |
| High[b] | 1,553 | 55·4 |
| **Current daily smoking** | | |
| No | 2,598 | 88·1 |
| Yes | 351 | 11·9 |
| **Alcohol consumption frequency** | | |
| Never to ≤monthly | 998 | 33·7 |
| 2-4/month or 2-3/week | 1,765 | 59·6 |
| ≥4/week | 198 | 6·7 |
| **Hospitalisation last 12 months** | | |
| No | 2,588 | 88·0 |
| Yes | 353 | 12·0 |
| **Diabetes mellitus**[c] | | |
| No | 2,674 | 94·0 |
| Yes | 170 | 6·0 |
| **Crohn`s disease/ulcerative colitis** | | |
| No | 2,831 | 97·9 |
| Yes | 60 | 2·1 |
| **Travel abroad past 12 months**[d] | | |
| No | 1,267 | 42·6 |
| Greece and Asia | 502 | 16·9 |
| All other countries | 1,171 | 39·4 |

[a] >College/university degree
[b] ≥551000 NOK (€ 53,767/year as per January 2021)
[c] 20 participants who answered "Yes, previously" were excluded
[d] Travelled outside the Nordic countries >1 week duration in the past 12 months.

**Supplementary Table 2**. Analysis of the significant variables in the multivariable model according to *Klebsiella* species

| | ***K. pneumoniae sensu stricto*, Kp1 (n=303)** | | **p-value[a]** | ***K. quasipneumoniae* subsp. *quasipneumoniae*, Kp2 (n=31)** | | **p-value[a]** | ***K. variicola* subsp. *variicola*, Kp3 (n=134)** | | **p-value[a]** |
|---|---|---|---|---|---|---|---|---|---|
| | **% (n) positive** | **% (n) negative** | | **% (n) positive** | **% (n) negative** | | **% (n) positive** | **% (n) negative** | |
| **Age (years)** | | | 0·052 | | | 0·003 | | | 0·435 |
| 40-49 | 6·1 (21) | 93·9 (323) | | 0·3 (1) | 99·7 (343) | | 4·1 (14) | 95·9 (330) | |
| 50-59 | 10·1 (44) | 89·9 (391) | | 2·1 (9) | 97·9 (426) | | 3·2 (14) | 96·8 (421) | |
| 60-69 | 11·2 (144) | 88·8 (1,142) | | 0·5 (6) | 99·5 (1,280) | | 5·1 (65) | 94·9 (1,221) | |
| 70-84 | 10·3 (94) | 89·7 (816) | | 1·6 (15) | 98·4 (895) | | 4·5 (41) | 95·5 (869) | |
| **Travel abroad past 12 months[b]** | | | 0·018 | | | 0·892 | | | 0·654 |
| No | 9·2 (116) | 90·8 (1,151) | | 1·0 (13) | 99·0 (1,254) | | 4·7 (59) | 95·3 (1,208) | |
| Greece or Asia | 13·5 (68) | 86·5 (434) | | 1·2 (6) | 98·8 (496) | | 5·2 (26) | 94·8 (476) | |
| All other countries | 9·6 (113) | 90·4 (1,058) | | 0·9 (11) | 99·1 (1,160) | | 4·2 (49) | 95·8 (1,122) | |
| **Crohn's disease/ulcerative colitis** | | | 0·038 | | | 0·431 | | | 0·137 |
| No | 10·1 (286) | 89·9 (2,545) | | 1·0 (29) | 99·0 (2,802) | | 4·3 (123) | 95·7 (2,708) | |
| Yes | 18·3 (11) | 81·7 (49) | | 0·0 (0) | 100 (60) | | 8·3 (5) | 91·7 (55) | |
| **Proton pump inhibitors last 6 m[c]** | | | 0·013 | | | 0·053 | | | 0·008 |
| No | 9·7 (255) | 90·3 (2,377) | | 0·9 (24) | 99·1 (2,608) | | 4·1 (109) | 95·9 (2,523) | |
| Yes | 14·0 (48) | 86·0 (295) | | 2·0 (7) | 98·0 (336) | | 7·3 (25) | 92·7 (318) | |
| **NSAIDs last 6 months[d]** | | | 0·469 | | | 0·020 | | | 0·157 |
| No | 10·0 (255) | 90·0 (2,290) | | 0·9 (22) | 99·1 (2,523) | | 4·3 (109) | 95·7 (2,436) | |
| Yes | 11·2 (48) | 88·8 (382) | | 2·1 (9) | 97·9 (421) | | 5·8 (25) | 94·2 (405) | |
| **Antibiotic systemic use last 1 m[e]** | | | 0·004 | | | 0·275 | | | 0·054 |
| No | 9·9 (283) | 90·1 (2,583) | | 1·1 (31) | 98·9 (2,835) | | 4·4 (125) | 95·6 (2,741) | |
| Yes | 18·3 (20) | 81·7 (89) | | 0·0 (0) | 100 (109) | | 8·3 (9) | 91·7 (100) | |

m, months; NSAIDs, nonsteroidal anti-inflammatory drugs; drug use according to the Norwegian Prescription Database.
[a] Determined using the Chi-square test.
[b] Travelled outside the Nordic countries >1 week duration in the past 12 months.
[c] A02BC, drugs used for peptic ulcer and gastro-oesophageal reflux disease.
[d] M01, anti-inflammatory and anti-rheumatic products (non-steroids), anti-inflammatory/anti-rheumatic agents in combination and specific anti-rheumatic agents.
[e] J01, A07AA09, P01AB01, antibacterials for systemic use, intestinal antiinfectives and nitroimidazole derivates used as antiprotozoals (metronidazole).

**Supplementary Table 3. ->** provided as Excel table

**Supplementary Table 4**. Matrix showing SNP differences among nine *K. pneumoniae sensu stricto* ST35 isolates

| | T7-208 | T7-272 | T7-276 | T7-290 | T7-330 | T7-358 | T7-360 | T7-396 | T7-479 |
|---|---|---|---|---|---|---|---|---|---|
| **T7-208** | | 311 | 310 | 310 | 310 | 312 | 310 | 309 | 272 |
| **T7-272** | 311 | | 1 | 1 | 4 | 1 | 0 | 2 | 309 |
| **T7-276** | 310 | 1 | | 0 | 1 | 2 | 0 | 1 | 308 |
| **T7-290** | 310 | 1 | 0 | | 0 | 2 | 0 | 1 | 308 |
| **T7-330** | 310 | 4 | 1 | 0 | | 5 | 1 | 1 | 311 |
| **T7-358** | 312 | 1 | 2 | 2 | 5 | | 0 | 3 | 310 |
| **T7-360** | 310 | 0 | 0 | 0 | 1 | 0 | | 1 | 308 |
| **T7-396** | 309 | 2 | 1 | 1 | 1 | 3 | 1 | | 309 |
| **T7-479** | 272 | 309 | 308 | 308 | 311 | 310 | 308 | 309 | |

**Supplementary Table 5**. Matrix showing SNP differences among five *K. pneumoniae sensu stricto* ST25 isolates

| | T7-067 | T7-127 | T7-161 | T7-308 | T7-451 |
|---|---|---|---|---|---|
| **T7-067** | | 30 | 3,066 | 29 | 29 |
| **T7-127** | 30 | | 3,045 | 7 | 7 |
| **T7-161** | 3,066 | 3,045 | | 3,044 | 3,044 |
| **T7-308** | 29 | 7 | 3,044 | | 8 |
| **T7-451** | 29 | 7 | 3,044 | 8 | |

**Supplementary Table 6.** SNP ranges within sequence types (ST) with more or equal than five *K. pneumoniae sensu stricto* isolates

| Sequence Type | No. of genomes | SNP range |
|---|---|---|
| **ST10** | 7 | 70-208 |
| **ST14** | 7 | 22-3,897 |
| **ST20** | 15 | 119-8,278 |
| **ST25** | 5 | 7-3,066 |
| **ST26** | 13 | 39-4,885 |
| **ST35** | 9 | 0-312 |
| **ST37** | 9 | 757-16,055 |
| **ST45** | 8 | 68-3,348 |
| **ST253** | 5 | 107-6,508 |
| **ST461** | 8 | 24-4,263 |
| **ST641** | 7 | 122-162 |
| **ST643** | 5 | 55-96 |
| **ST681** | 7 | 60-200 |
| **ST1423** | 5 | 115-396 |
| **ST1562** | 6 | 39-11,993 |
| **ST2004** | 5 | 13-23 |
| **ST2386** | 9 | 41-78 |

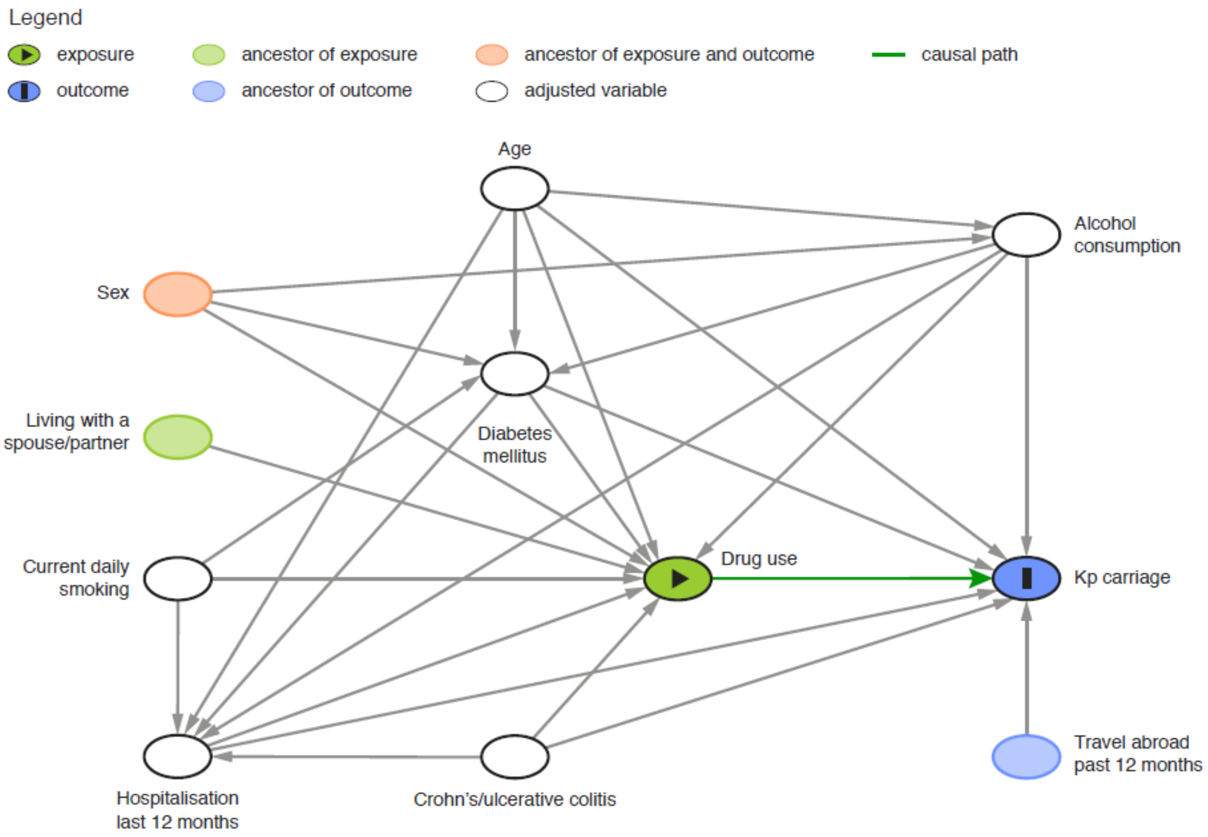


**Supplementary Figure 1**. Directed acyclic graph (DAG) illustrating causal relationships between drug use (exposure), *K. pneumoniae* (Kp) faecal carriage (outcome) and relevant covariates. DAG was used for selection of the multivariable logistic regression model. White variables are those adjusted for. Even if sex is an ancestor of outcome, there is no biasing path implying that sex should not be included in the model. The drug use variables include proton pump inhibitors, non-steroidal anti-inflammatory drugs, metformin and thyroid hormones used the last six months, and antibacterials for systemic use the last one month.

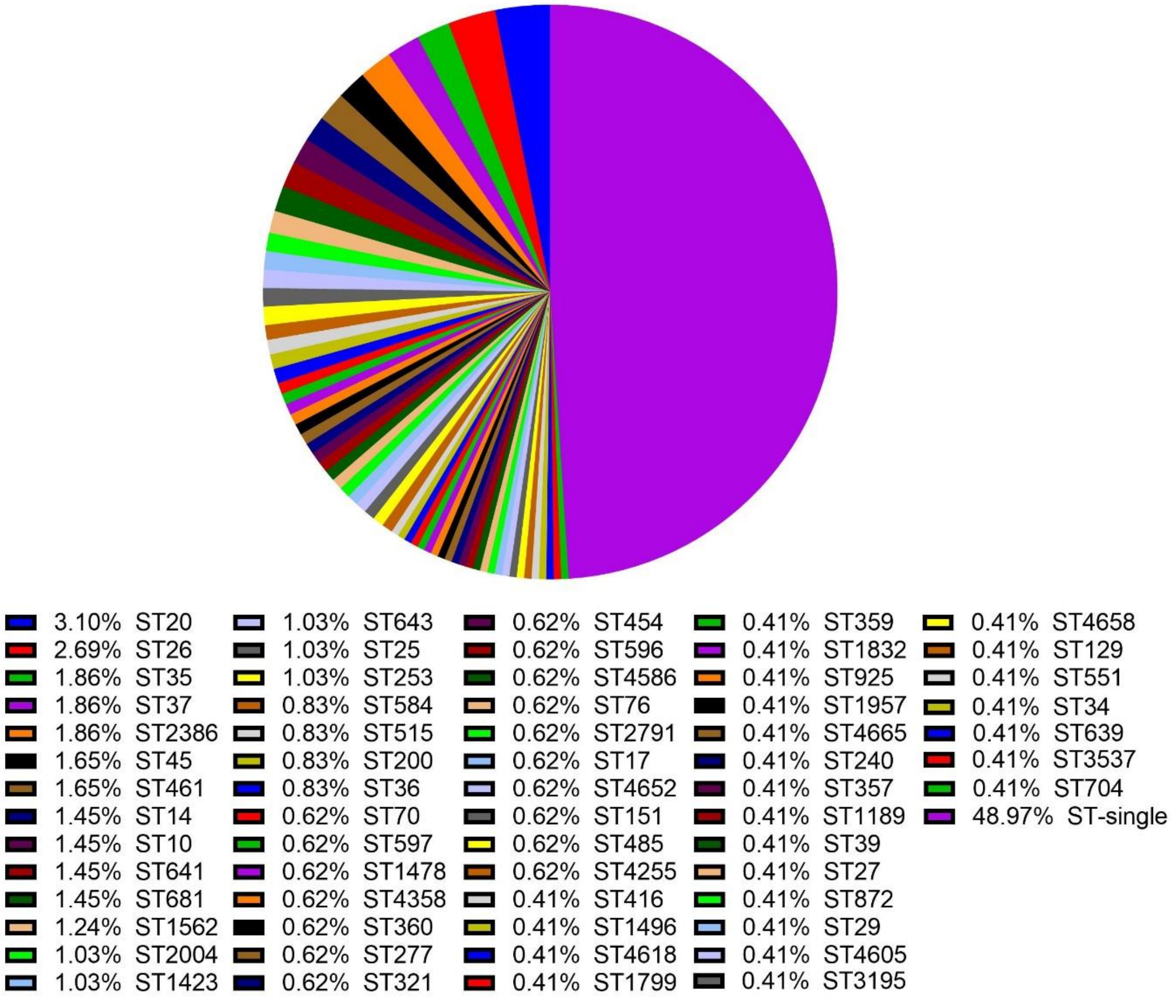


**Supplementary Figure 2**. Multilocus sequence type diversity in 484 *K. pneumoniae* species complex isolates including 300 different sequence types (STs). ST and proportion (%) is indicated in the colour legend. Unique STs represented by one isolate is grouped in the ST-single (large violet) area. In total, 32% (96 of 300) of STs identified were novel.

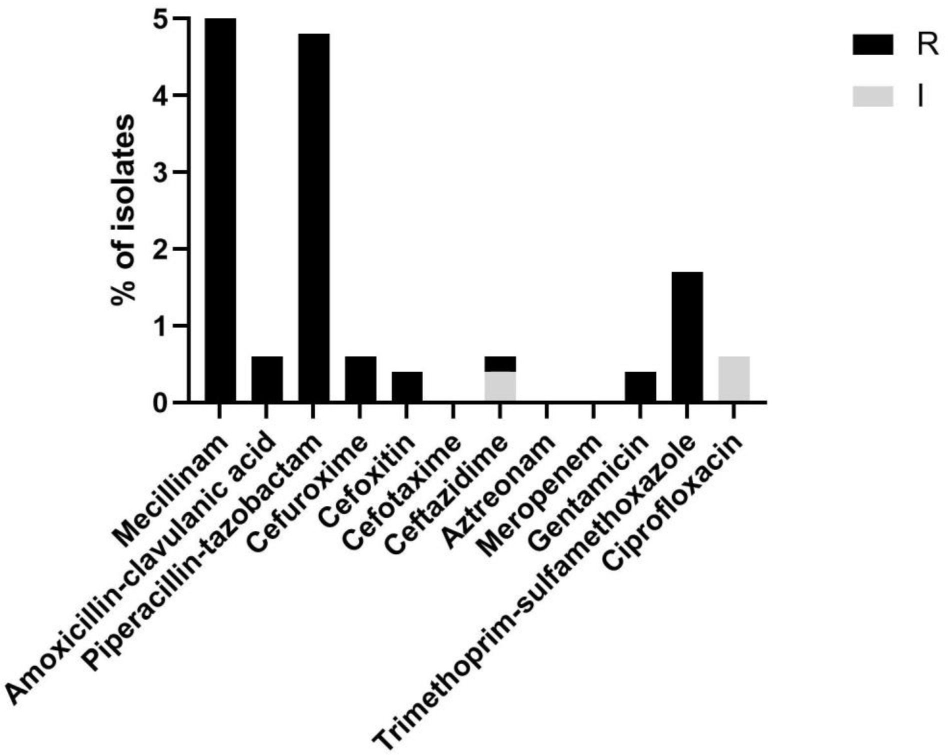


**Supplementary Figure 3**. Prevalence of phenotypic antibiotic resistance (R) and susceptible, increased exposure (I) among 484 *K. pneumoniae* species complex isolates.

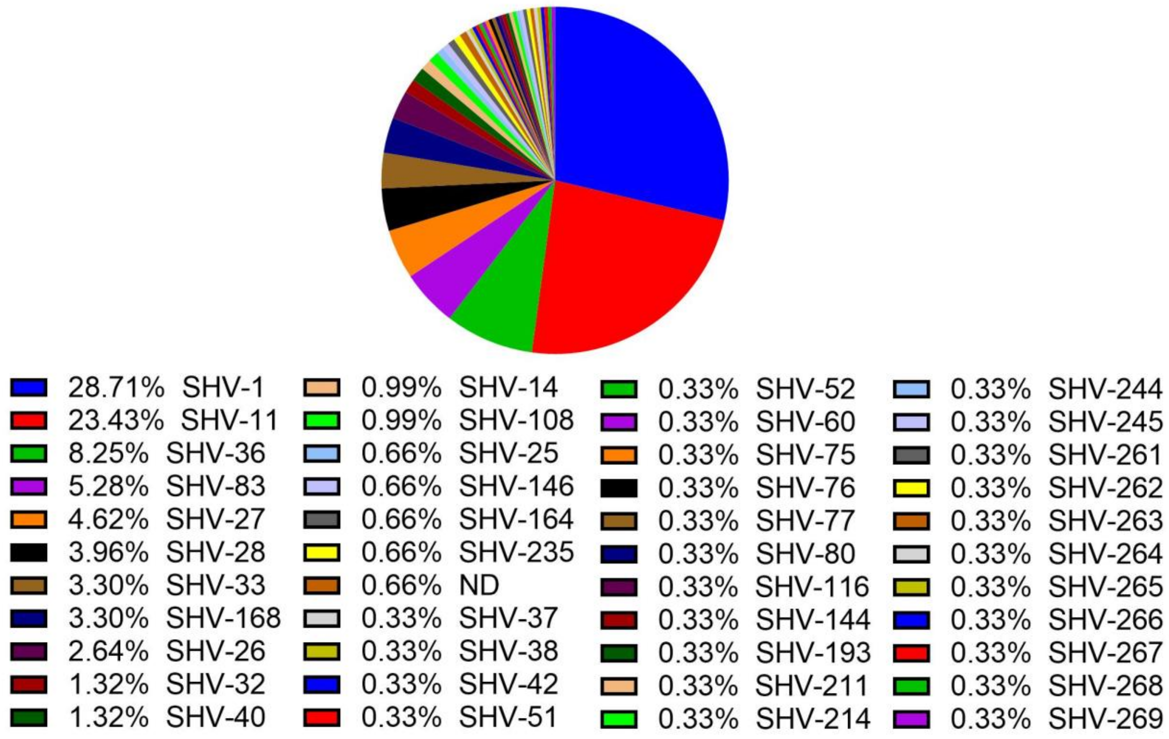


**Supplementary Figure 4.** SHV β-lactamase diversity among *K. pneumoniae sensu stricto* (Kp1) isolates (n=303). SHV-variant and proportion (%) is indicated in the legend. Two isolates with either a deleted *bla*$_{SHV}$ gene or a *bla*$_{SHV}$ gene with a premature stop codon are included and labelled ND.

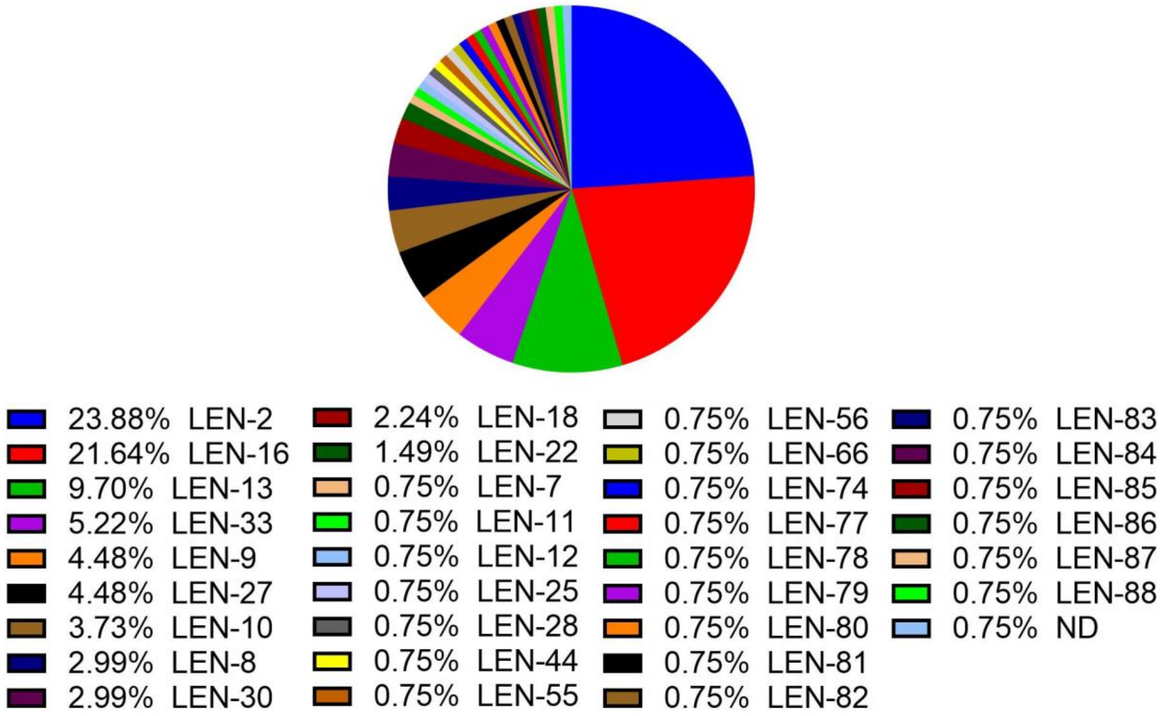


**Supplementary Figure 5**. LEN β-lactamase diversity among *K. variicola* subsp. *variicola* (Kp3) isolates (n=134). LEN-variant and proportion (%) is indicated in the legend. One isolate with a deleted $bla_{\text{LEN}}$ gene is included and labelled ND.

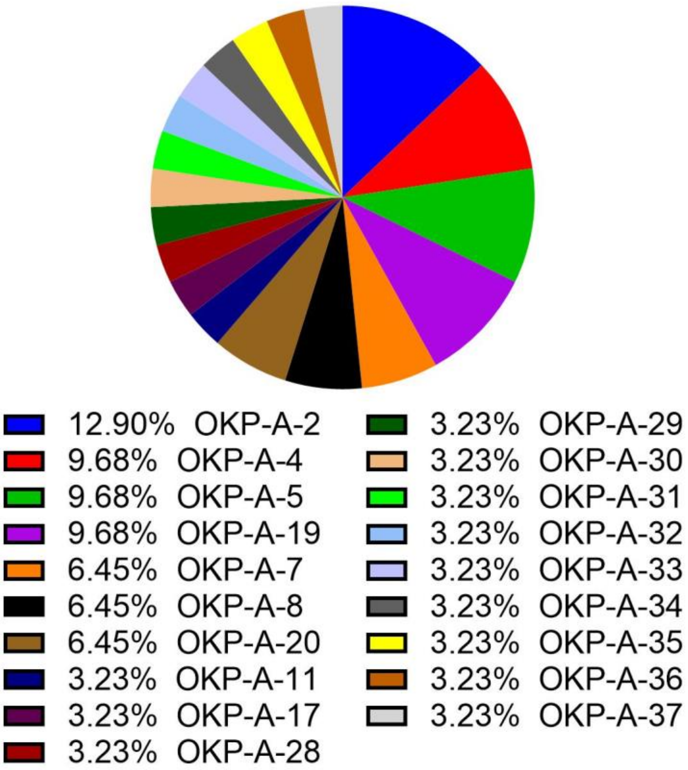


**Supplementary Figure 6**. OKP-A β-lactamase diversity among *K. quasipneumoniae* subsp. *quasipneumoniae* (Kp2) isolates (n=31). OKP-A-variant and proportion (%) is indicated in the legend.

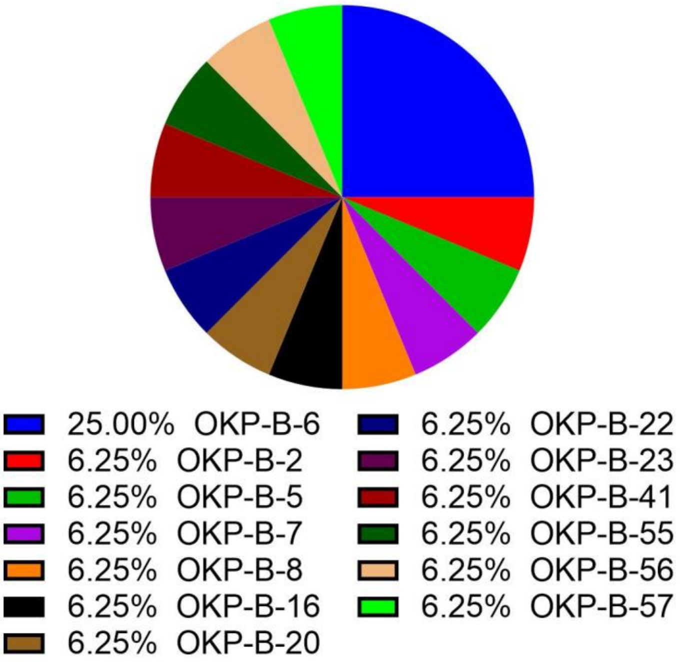


**Supplementary Figure 7**. OKP-B β-lactamase diversity among *K. quasipneumoniae* subsp. *similipneumoniae* (Kp4) isolates (n=16). OKP-B-variant and proportion (%) is indicated in the legend.

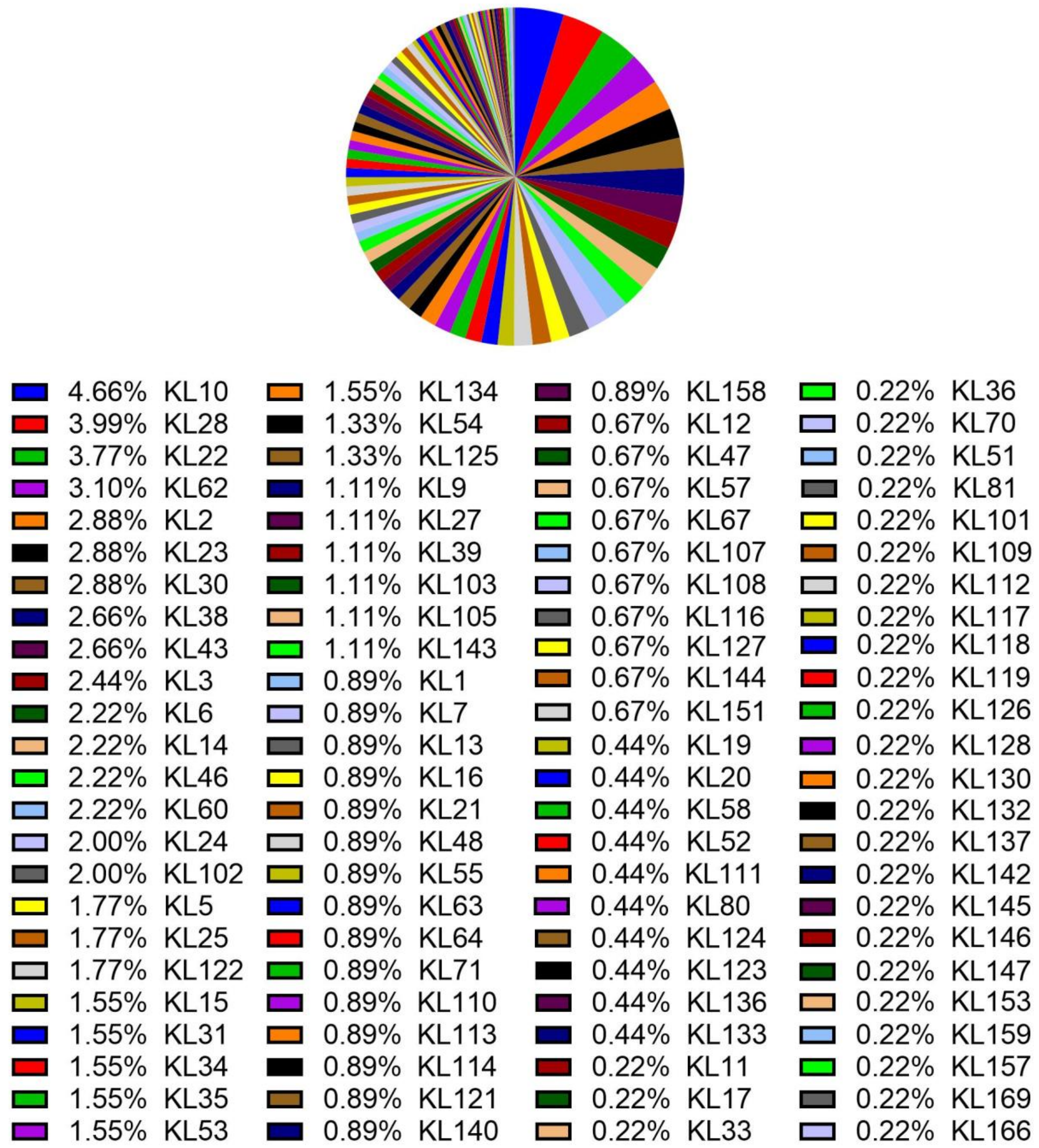


**Supplementary Figure 8**: Capsule locus (KL) type diversity among 451 *K. pneumoniae* species complex isolates with a defined KL type and proportion (%) indicated in the legend.[1]

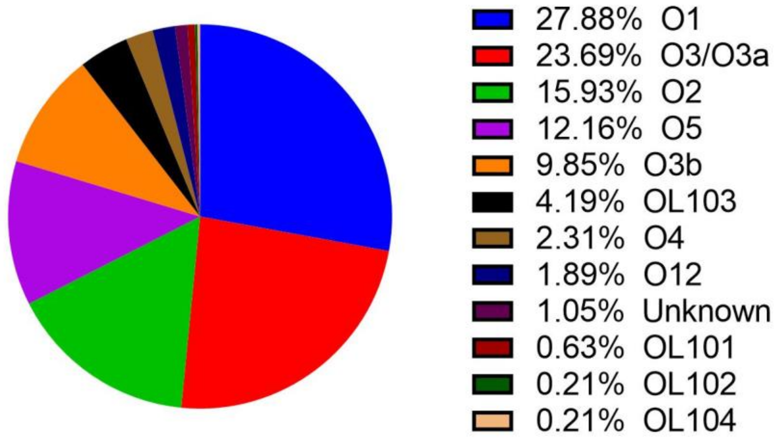


**Supplementary Figure 9**. O type diversity among 477 *K. pneumoniae* species complex isolates with a defined O type and proportion (%) is indicated in the legend. Unknown represents when only one of the two additional genes (*wbbY* and *wbbZ*) can be found, the result is ambiguous and can either be O1 or O2.[1]
